# Multimodal Artificial Intelligence Predicts Pathological Response and Prognosis in HER2-Positive Early Breast Cancer Treated with Chemotherapy De-escalation Strategies: Analyses from PHERGain and PHERGain-2 Trials

**DOI:** 10.64898/2026.10.05.26364736

**Authors:** Manuel Ruiz Borrego, Javier Cortés, Agostina Stradella, Begoña Bermejo, Santiago Escrivá-de-Romaní, Cristina Reboredo, Cinta Albacar, Laia Garrigós, Sherko Kümmel, Marco Colleoni, Geraldine Gebhart, Florence Dalenc, Khaldoun Kerrou, Sofia Braga, Peter Schmid, Frederik Marmé, Serena Di Cosimo, Giulia Notini, Elena Martínez-García, Ana Amaya-Garrido, Daniel Alcalá-López, Leonardo Mina, Jose Rodriguez-Morató, Cerise Tang, Dhruva Biswas, Bartosz Machura, Jan Witowski, Krzysztof J. Geras, Jungkyu Park, Mario Mancino, Jose Manuel Pérez-García, Antonio Llombart-Cussac

## Abstract

**Background:** Chemotherapy (CT) de-escalation strategies based on dual HER2 blockade with trastuzumab and pertuzumab (HP) have shown promising efficacy in HER2-positive (HER2+) early breast cancer (EBC), but biomarkers to guide patient selection are lacking. The Ataraxis Breast platform (ATX) uses artificial intelligence (AI) models integrating hematoxylin-eosin (HCE) whole-slide images and clinical variables to predict pathological complete response (pCR) and long-term outcomes. We evaluated ATX in the phase II PHERGain (NCT03161353) and PHERGain-2 (NCT04733118) trials.

**Methods:** The ATX NEO model was assessed for pCR prediction in PHERGain (n=306 evaluable patients) and PHERGain-2 (n=383 evaluable patients). The ATX RISK model was evaluated for prognosis in PHERGain (n=309 evaluable patients). Prespecified cutoffs classified patients into low-, medium-, and high-probability groups for pCR and low-or high-risk groups for recurrence. Associations with pCR were assessed using logistic regression. Prognostic performance was evaluated using Cox proportional hazards models, Kaplan-Meier estimates, and Harrell’s C-index.

**Results:** Overall pCR rates were 40.2% in PHERGain and 60.3% in PHERGain-2. Continuous NEO scores were significantly associated with pCR in both PHERGain (odds ratio [OR], 1.28; 95% CI, 1.11-1.48; P<0.001) and PHERGain-2 (OR, 1.30; 95% CI, 1.13-1.52; P<0.001). Using prespecified cutoffs, pCR rates increased across low-, medium-, and high-probability groups (19.2%, 41.3%, and 48.3% in PHERGain; 29.5%, 61.6%, and 70.1% in PHERGain-2). In PHERGain, the RISK score was significantly associated with event-free survival (hazard ratio per 1-SD increase, 1.56; 95% CI, 1.17-2.08; P=0.002; C-index, 0.66). Five-year event-free survival was 93.9% in the low-risk group and 83.1% in the high-risk group.

**Conclusions:** NEO score was consistently associated with pCR across two independent HER2+ EBC trials, while RISK score provided prognostic information for long-term outcomes in PHERGain. Prespecified score categories identified clinically distinct response and risk groups. These findings support further evaluation of AI-based biomarkers to refine patient selection for CT de-escalation strategies in HER2+ EBC.

## INTRODUCTION

Neoadjuvant systemic therapy for HER2-positive (HER2+) early breast cancer (EBC) has changed substantially over the past decade, as dual HER2 blockade with trastuzumab and pertuzumab (HP) plus chemotherapy (CT) has become a widely adopted standard, supported by increased rates of pathologic complete response (pCR) in the neoadjuvant setting and improved invasive disease-free survival (iDFS) in selected higher-risk populations ^1,2^. However, this approach exposes patients to substantial acute and cumulative toxicity, fueling interest in strategies that can preserve excellent outcomes while reducing or omitting CT ^1,3^. These competing realities have motivated response-adapted de-escalation strategies seeking to reduce or omit CT without compromising long-term disease control ^4–6^.

PHERGain evaluated an 18F-FDG-PET- and pCR-adapted strategy in stage I–IIIA HER2+ EBC, allowing approximately one third of patients to avoid chemotherapy with favorable 3-year iDFS ^4,5^. PHERGain-2 further explored a CT-free neoadjuvant HP approach in a favorable-risk population with centrally confirmed HER2 immunohistochemistry (IHC) 3+, node-negative disease, and small tumors, achieving high pCR rates and improved quality of life, although long-term efficacy remains under evaluation ^6^. Although these trials showed that de-escalated or CT-free treatment is feasible in selected patients, they also underscored the persistent need for biomarkers that can prospectively enrich for favorable biology and reliably identify candidates for CT omission.

Despite advances in molecular characterization, no biomarker has yet achieved widespread clinical adoption for prospectively selecting patients with HER2+ EBC for CT omission or substantial de-escalation in the neoadjuvant setting. Clinicopathologic factors and molecular tools can provide prognostic and predictive information but still have imperfect ability to pinpoint those most likely to achieve pCR with HER2-directed therapy alone or to sustain long-term disease control after de-escalation ^7–9^. Advances in artificial intelligence (AI)-based computational pathology have enabled the extraction of high-dimensional morphologic and spatial features from routine hematoxylin-and-eosin (HCE) whole-slide images, potentially capturing clinically relevant tumor and microenvironmental characteristics beyond conventional histopathologic assessment, using widely available tissue material without the need for additional molecular testing ^10–13^. This translational study of the PHERGain and PHERGain-2 trials was therefore conceived to evaluate whether AI–based analysis of routine HCE whole-slide images, integrated with standard clinical variables, could generate clinically relevant predictive and prognostic information in this setting.

Building on this rationale, the Ataraxis Breast platform (ATX) generates two conceptually distinct outputs by integrating digital pathology with clinical inputs: NEO, which estimates the probability of pCR after neoadjuvant therapy, and RISK, which estimates recurrence risk ^13^. The present retrospective study has two primary objectives: first, to determine whether the NEO score predicts pCR in PHERGain and PHERGain-2; and second, to assess whether the RISK score is prognostic for 5-year outcomes in the PHERGain trial, as long-term outcome data from PHERGain-2 are not yet available.

## METHODS

### Study design and participants

PHERGain (NCT03161353) was a randomized phase II trial enrolling patients with centrally confirmed stage I–IIIA HER2+ EBC and an ^18^F-FDG-PET-evaluable lesion. Patients received either neoadjuvant TCHP or an adaptive strategy initially comprising HP, with subsequent treatment determined by early ^18^F-FDG-PET response and pCR ^4,5^. At this updated analysis (data cutoff, November 4, 2024), the median duration of follow-up was 5.6 years (range, 0.4 to 7.3). PHERGain-2 (NCT04733118) was a single-arm phase II trial evaluating neoadjuvant trastuzumab and pertuzumab without chemotherapy in patients with centrally confirmed HER2 IHC 3+, node-negative tumors measuring 5–30 mm ^6^. Full eligibility criteria and treatment procedures have been previously reported. PHERGain-2 follow-up remains ongoing.

### AI platform and biomarker definitions

NEO is a multimodal AI model that integrates information extracted from baseline HCE whole-slide images with clinical variables to estimate the probability of pCR. The model uses pathology foundation models and multiple-instance learning to derive image-based features and inferred gene-expression representations, which are combined with baseline clinicopathologic variables. NEO was developed and externally validated across multiple independent cohorts. Prespecified thresholds classified patients into low (<0.30), medium (0.30–0.55), and high (>0.55) predicted probability of pCR categories.

RISK is a multimodal AI model that integrates HCE whole-slide images and clinical variables to estimate 5-year recurrence risk. The model combines image-derived features, inferred gene-expression representations, and clinicopathologic variables within an ensemble survival framework trained and validated across multiple independent cohorts. A prespecified threshold classified patients as having low (<0.20) or high (≥0.20) predicted 5-year recurrence risk. Neither PHERGain nor PHERGain-2 data were used for model development or training.

Detailed model architecture, development, training, validation procedures, and performance metrics are provided in the **Supplementary Methods**.

### Endpoints

The primary predictive endpoint was pCR, defined as the absence of residual invasive cancer in the breast and axillary lymph nodes at definitive surgery (ypT0/is ypN0), as assessed locally. The association between the baseline NEO score and pCR was evaluated separately in PHERGain and PHERGain-2, both using the continuous score and prespecified low-, medium-, and high-probability groups. The definition of pCR slightly differed between PHERGain and PHERGain-2, with ypN0(i+) or ypN0(mol+) considered node-negative in PHERGain but residual disease in PHERGain-2; however, only a small number of patients (n=4) in each trial were affected by this distinction.

The primary prognostic endpoint for evaluation of the baseline RISK score in PHERGain was event-free survival (EFS), defined as the time from randomization to disease progression during neoadjuvant treatment, invasive ipsilateral breast tumor recurrence, locoregional invasive recurrence, distant recurrence, contralateral invasive breast cancer, a second primary non-breast invasive cancer, or death from any cause, whichever occurred first.

Additional prognostic endpoints were invasive disease-free survival (iDFS), invasive breast cancer-free survival (IBCFS), distant disease-free survival (DDFS), and overall survival (OS). iDFS, IBCFS, and DDFS were measured from the date of definitive surgery and evaluated among patients who underwent protocol-defined surgery. EFS and OS were measured from the date of randomization and evaluated among all patients with a valid baseline RISK score. Endpoint definitions followed the Standardized Definitions for Efficacy End Points in Adjuvant Breast Cancer Trials (STEEP) criteria. OS was defined as the time from randomization to death from any cause. Patients without an event were censored at the date of their last available disease assessment or contact.

All biomarker endpoints were assessed retrospectively. The analyses of the NEO score evaluated its association with pathological response, whereas the analyses of the RISK score evaluated its prognostic association with long-term clinical outcomes.

### Statistical Analysis

The association between the continuous NEO score and pCR was assessed by univariable logistic regression separately in each trial. Prespecified cutoffs defined low-, medium-, and high-probability groups, and pCR rates were compared across these groups using odds-ratio contrasts, including a prespecified comparison between the high- and the low-probability groups. The incremental value of NEO was evaluated using nested logistic models comprising NEO alone, clinicopathological variables alone, and both combined. Discrimination was assessed using apparent and bootstrap optimism-corrected AUCs, and added value was tested by likelihood-ratio tests.

The association between the continuous RISK score and clinical outcomes was evaluated using Cox proportional-hazards models with hazard ratios (HRs) reported per 1-standard-deviation increase in the score. Kaplan–Meier methods estimated time-to-event outcomes, and model discrimination was assessed using Harrell’s concordance index (C-Index).

## RESULTS

### Patients and AI-evaluable cohorts

Of 356 patients randomized to Groups A and B in PHERGain, 335 (94.1%) had a baseline core needle biopsy available, and 306 (85.9%) yielded a valid NEO score after quality control of the whole-slide images. A valid baseline RISK score was available for 309 (86.8%) patients, including the 306 patients with a valid NEO score and three additional patients whose slides yielded a RISK but not a NEO score. Of these, 290 (81.5% of the randomized population) underwent definitive surgery and were included in the time-to-event analyses (iDFS, IBCFS, and DDFS).

In PHERGain-2, 392 of 396 (98.9%) patients had a baseline core needle biopsy available, and 383 (96.7%) yielded a valid NEO score after quality control of the whole-slide images. RISK scores were not generated for PHERGain-2 because long-term outcome data are not yet available.

Overall, 689 patients across the two trials had NEO scores and were included in the predictive analyses, with each trial analyzed separately. The overall pCR rate was 40.2% (95% confidence interval [CI], 34.7 to 45.9) in PHERGain and 60.3% (95% CI, 55.2 to 65.2) in PHERGain-2. Baseline characteristics of the NEO-analyzable populations are summarized in **Table 1**.

**Table 1.** Patient characteristics.

|  | **PHERGain**<br>**(N=309)** | **PHERGain-2**<br>**(N=383)** |
| --- | --- | --- |
| **Age in years**, median (range) | 50.0 (25.0–77.0) | 56.3 (25.5–85.9) |
| **Menopausal status** |  |  |
| Premenopausal | 156 (50.5%) | 151 (39.4%) |
| Postmenopausal | 153 (49.5%) | 230 (60.1%) |
| Missing | – | 2 (0.5%) |
| **Clinical tumor stage (cT)** |  |  |
| T1 | 46 (14.9%) | 246 (64.2%) |
| T2 | 210 (67.9%) | 137 (35.8%) |
| T3 | 53 (17.2%) | 0 (0.0%) |
| **Clinical nodal stage (cN)** |  |  |
| N0 | 156 (50.5%) | 382 (99.7%) |
| N1 | 135 (43.7%) | 1 (0.3%) |
| N2 | 18 (5.8%) | 0 (0.0%) |
| **Clinical stage** |  |  |
| I | 31 (10.10%) | 245 (64.0%) |
| II | 230 (74.4%) | 138 (36.0%) |
| III | 48 (15.5%) | 0 (0.0%) |
| **Histological grade** |  |  |
| G1-G2 | 121 (39.2%) | 200 (52.9%) |
| G3 | 140 (45.3%) | 127 (33.6%) |
| Gx | 48 (15.5%) | 51 (13.5%) |
| Missing | 0 | 5 |
| **Hormone receptor status** |  |  |
| Negative | 104 (33.7%) | 104 (27.2%) |
| Positive | 205 (66.3%) | 279 (72.8%) |
| **Pathological complete response** |  |  |
| No | 186 (60.2%) | 152 (39.7%) |
| Yes | 123 (39.8%) | 231 (60.3%) |
| **NEO score** |  |  |
| Low | 52 (17.0%) | 44 (11.5%) |
| Medium | 138 (45.1%) | 232 (60.6%) |
| High | 116 (37.9%) | 107 (27.9%) |
| Missing | 3 | – |
| **RISK score** |  |  |
| Low | 158 (51.1%) | – |
| High | 151 (48.9%) | – |

Percentages may not sum to 100% owing to rounding.

### Association between NEO score and pCR

The NEO model was evaluated both as a continuous score and according to prespecified low-, medium-, and high-probability groups for pCR (**Table 2**). In PHERGain, each 0.1-unit increase in the continuous NEO score was associated with higher odds of achieving pCR in univariable analysis (odds ratio, 1.28; 95% CI, 1.11 to 1.48; P<0.001). This association remained significant after multivariable adjustment for treatment group and hormone receptor status (adjusted OR, 1.24; 95% CI, 1.02 to 1.52; P=0.033) (**Table 2 and Table S1**). In PHERGain-2, each 0.1-unit increase in the NEO score was similarly associated with pCR in univariable analysis (OR, 1.30; 95% CI, 1.13 to 1.52; P<0.001) and after adjustment for hormone receptor status (adjusted OR, 1.43; 95% CI, 1.18 to 1.74; P<0.001) **(Table 2 and Table S1)**. Receiver-operating-characteristic (ROC) analyses yielded areas under the curve (AUCs) of 0.613 (95% CI, 0.549 to 0.677) in PHERGain and 0.618 (95% CI, 0.560 to 0.676) in PHERGain-2 (**Figure S1**). NEO improved the clinical model in PHERGain and PHERGain-2, increasing optimism-corrected AUCs from 0.567 to 0.611 and from 0.518 to 0.592, respectively (both P<0.001; **Table S3**); clinical variables did not significantly improve NEO alone.

**Table 2.** NEO score association for pCR.

|  | UNIVARIABLE |  | MULTIVARIABLE |  |
| --- | --- | --- | --- | --- |
| Predictor | OR (95% CI) | p | OR (95% CI) | p |
| **PHERGain (n = 306 UV/302 MV)** |  |  |  |  |
| **Continuous, per 0.1 NEO** | 1.28 (1.11–1.48) | <0.001 | 1.24 (1.02–1.52) | 0.033 |
| **NEO medium vs low** | 2.96 (1.42–6.67) | 0.006 | 2.64 (1.25–6.04) | 0.015 |
| **NEO high vs low** | 3.92 (1.86–8.95) | <0.001 | 2.77 (1.13–7.17) | 0.030 |
| **PHERGain-2 (n = 383)** |  |  |  |  |
| **Continuous, per 0.1 NEO** | 1.30 (1.13–1.52) | <0.001 | 1.43 (1.18–1.74) | <0.001 |
| **NEO medium vs low** | 3.83 (1.94–7.95) | <0.001 | 3.99 (2.01–8.31) | <0.001 |
| **NEO high vs low** | 5.59 (2.64–12.39) | <0.001 | 6.85 (2.91–16.84) | <0.001 |

Odds ratios are per 0.1-unit increase in the continuous NEO score, or versus the low-probability group. Confidence intervals are profile likelihood. Multivariable models adjust for treatment arm and hormone receptor status (PHERGain) and for hormone receptor status (PHERGain-2).

Using the prespecified NEO cutoffs, 52 of 306 patients (17.0%) in PHERGain were classified in the low-probability group, 138 (45.1%) in the medium-probability group, and 116 (37.9%) in the high-probability group. pCR was observed in 19.2% (10/52; 95% CI, 9.6 to 32.5), 41.3% (57/138; 95% CI, 33.0 to 50.0), and 48.3% (56/116; 95% CI, 38.9 to 57.7) of patients, respectively (**Figure 1**). In the prespecified comparison between the high- and low-probability groups, patients in the high-probability group had higher odds of achieving pCR (OR, 3.92; 95% CI, 1.86–8.95; P<0.001). The medium-probability group also had higher odds of pCR compared with the low-probability group (OR, 2.96; 95% CI, 1.42–6.67; P=0.006). These associations persisted after adjustment for treatment group and hormone receptor status (**Table 2**).

**Figure 1.**
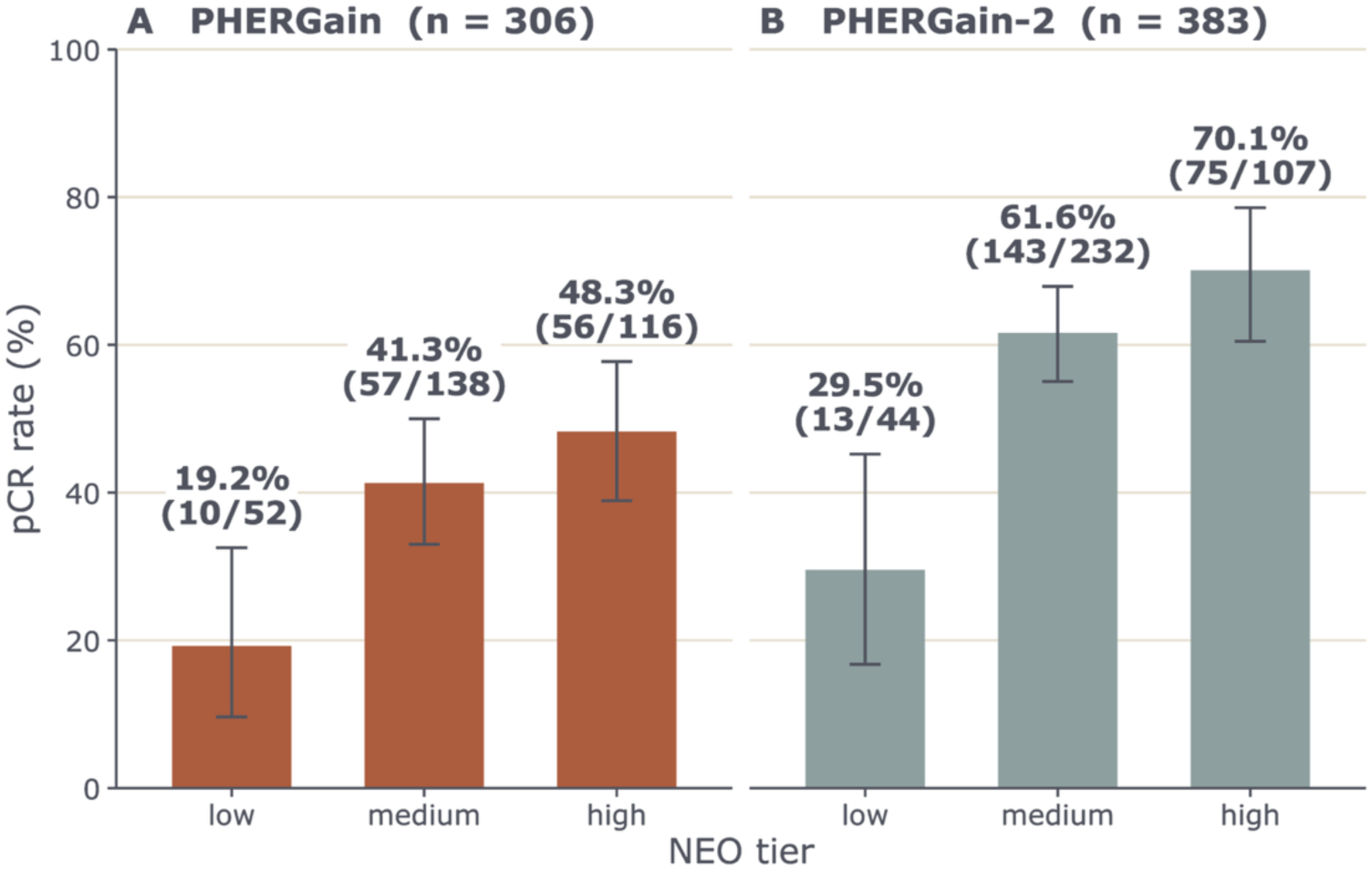
pCR rates according to NEO categories in PHERGain and PHERGain-2. Bar plots showing pCR rates across low, medium, and high baseline NEO tiers in PHERGain (A; n = 306) and PHERGain-2 (B; n = 383). Bars represent the proportion of patients achieving pCR within each NEO probability group, with percentages and responder counts indicated above each bar. Error bars denote 95% confidence intervals.

Among 383 patients in PHERGain-2, 44 (11.5%) were classified in the low-probability NEO group, 232 (60.6%) in the medium-probability group, and 107 (27.9%) in the high-probability group. pCR was observed in 29.5% (13/44; 95% CI, 16.8 to 45.2), 61.6% (143/232; 95% CI, 55.0 to 67.9), and 70.1% (75/107; 95% CI, 60.5 to 78.6) of patients, respectively. In the prespecified comparison between the high- and low-probability groups, patients in the high-probability group had higher odds of achieving pCR (OR, 5.59; 95% CI, 2.64–12.39; P<0.001). The medium-probability group also had higher odds of pCR compared with the low-probability group (OR, 3.83; 95% CI, 1.94–7.95; P<0.001). These associations persisted after adjustment for hormone receptor status (adjusted OR, 3.99; 95% CI, 2.01 to 8.31; P<0.001 for the medium-probability group; and adjusted OR, 6.85; 95% CI, 2.91 to 16.84; P<0.001 for the high-probability group; likelihood-ratio P<0.001 for the NEO group effect; **Table 2**).

### NEO performance across treatment groups and ^18^F-FDG-PET response subgroups in PHERGain

In PHERGain, the association between NEO probability groups and pCR was further explored across treatment groups and ^18^F-FDG-PET response subgroups in Group B (**Figure 2**). Greater NEO discrimination among patients receiving upfront TCHP than among those managed using the PET-adapted strategy was observed, with AUCs of 0.693 and 0.593, respectively. Within the adaptive-treatment group, the AUC was 0.559 among PET responders and 0.729 among ^18^F-FDG PET non-responders. However, subgroup sizes were limited, and the NEO-by-PET-response interaction was not statistically significant (p=0.097).

**Figure 2.**
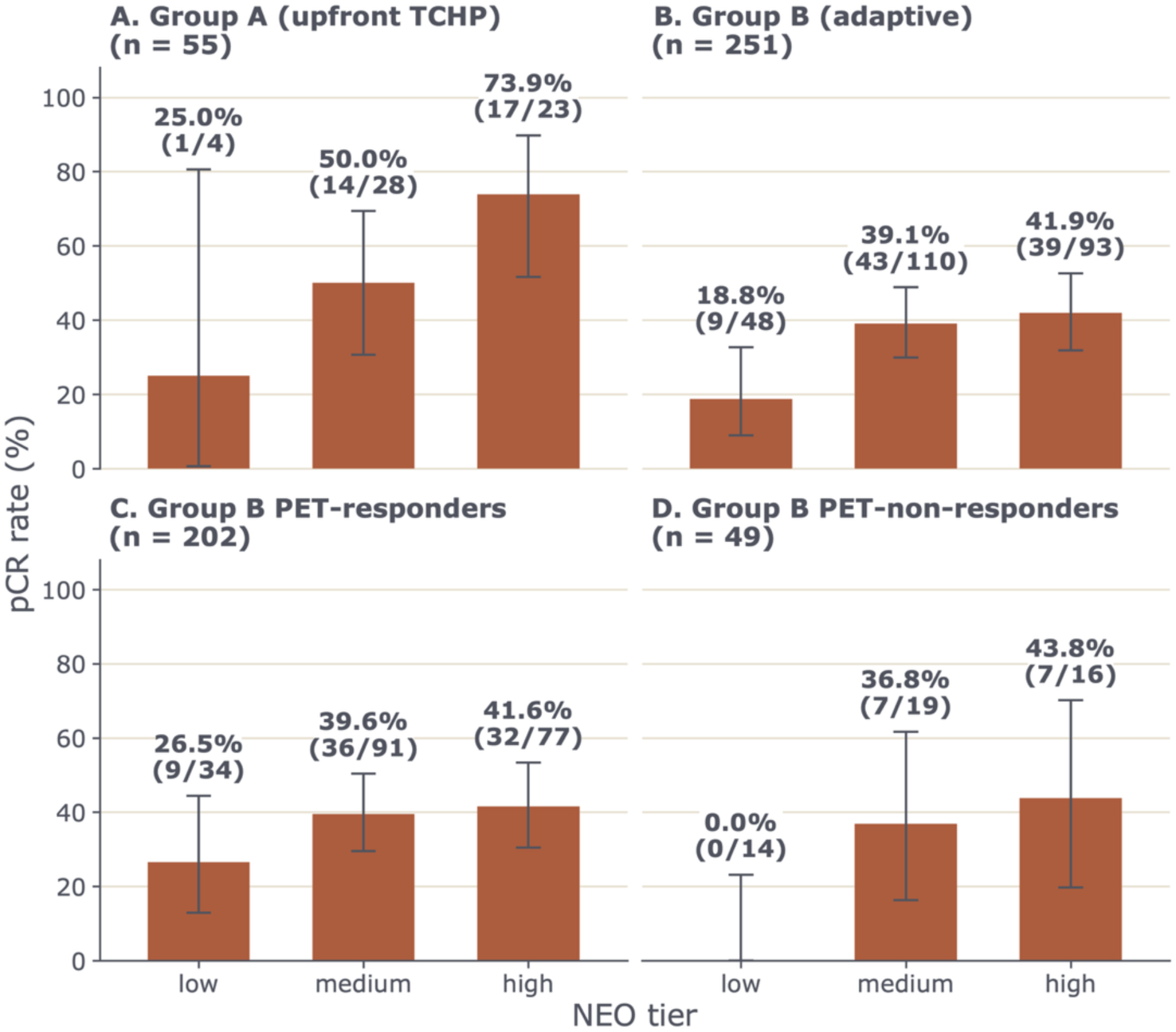
pCR rates according to baseline NEO probability groups across treatment groups and ^18^F-FDG-PET response status in PHERGain. Bar plots showing pCR rates stratified by baseline NEO probability groups (low, medium, and high) in (A) Group A, patients treated with upfront TCHP (n = 55); (B) Group B, patients managed with the adaptive treatment strategy (n = 251); (C) ^18^F-FDG**-**PET responders within Group B (n = 202); and (D) ^18^F-FDG**-**PET non-responders within Group B (n = 49). Bars represent the proportion of patients achieving pCR, with percentages and responder counts displayed above each bar. Error bars indicate 95% confidence intervals.

### Prognostic value of the RISK model in PHERGain

Prognostic analyses were performed in 309 patients from PHERGain with a valid baseline RISK score. Analysis of disease-free-survival endpoints (iDFS, IBCFS, and DDFS) were restricted to the 290 patients who underwent definitive surgery, as these endpoints were measured from the date of surgery. The continuous RISK score was significantly associated with EFS (HR per 1-standard deviation (SD) increase, 1.56; 95% CI, 1.17 to 2.08; P=0.002; Harrell’s C-index, 0.66) (**Table 3**). Based on the prespecified RISK score cutoff, 158 (51.1%) patients were classified as low risk and 151 (48.9%) as high risk, with 5-year EFS rates of 93.9% (95% CI, 90.2 to 97.9) and 83.1% (95% CI, 77.1 to 89.5), respectively (log-rank P=0.025; **Figure 3**).

**Figure 3.**
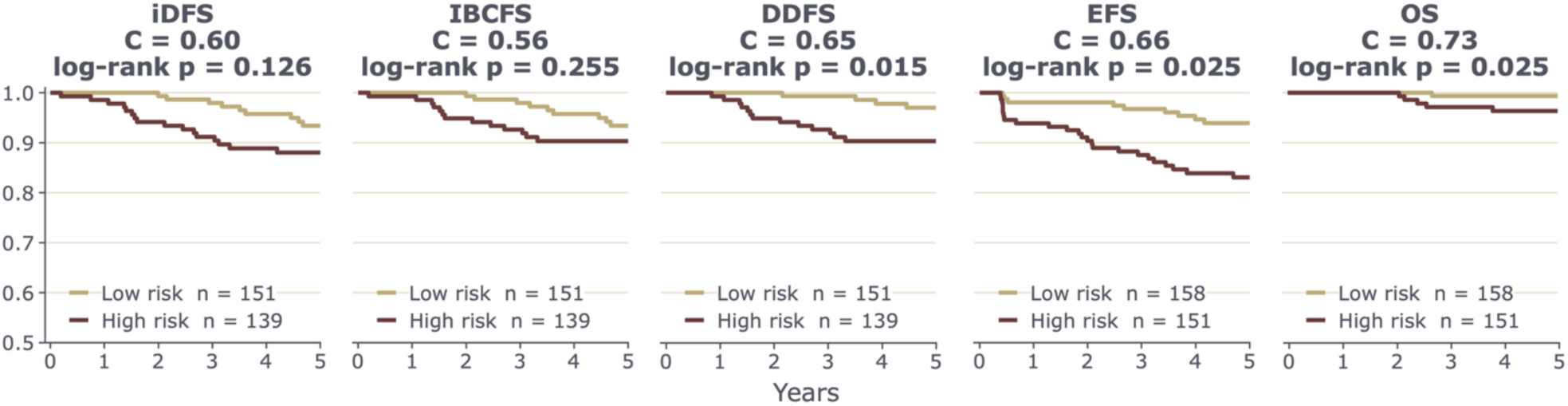
Kaplan–Meier estimates of time-to-event outcomes according to the RISK model in PHERGain. Kaplan–Meier estimates of invasive disease–free survival (iDFS), invasive breast cancer–free survival (IBCFS), distant disease–free survival (DDFS), event-free survival (EFS), and overall survival (OS) according to the Ataraxis RISK model in PHERGain. EFS and OS analyses included all 309 patients with a valid baseline RISK score, whereas iDFS, IBCFS, and DDFS analyses were restricted to the 290 patients who underwent definitive surgery. P values shown in each panel are log-rank tests comparing the prespecified low- and high-risk groups.

**Table 3.** Prognostic value of RISK score association to 5-year efficacy outcomes.

|  |  |  | Continuous RISK score |  |  | Prespecified RISK groups |  |  |
| --- | --- | --- | --- | --- | --- | --- | --- | --- |
|  |  |  | *Cox model, per 1-SD increase* |  |  | *Kaplan-Meier* |  |  |
| Endpoint | Patients analyzed | Events | HR / 1 SD (95% CI) | P | C-index | 5-y low risk % (95% CI) | 5-y high risk % (95% CI) | Log-rank P |
| iDFS | 290 | 26 | 1.22 (0.85–1.75) | 0.282 | 0.60 | 93.4 (89.3–97.7) | 88.0 (82.7–93.7) | 0.126 |
| IBCFS | 290 | 22 | 1.10 (0.73–1.65) | 0.661 | 0.56 | 93.4 (89.3–97.7) | 90.3 (85.5–95.5) | 0.255 |
| DDFS | 290 | 17 | 1.34 (0.87–2.06) | 0.185 | 0.65 | 97.0 (94.1–99.9) | 90.3 (85.5–95.5) | 0.015 |
| EFS | 309 | 37 | 1.56 (1.17–2.08) | 0.002 | 0.66 | 93.9 (90.2–97.9) | 83.1 (77.1–89.5) | 0.025 |
| OS | 309 | 8 | 1.72 (0.96–3.11) | 0.070 | 0.73 | 99.3 (98.1–100.0) | 96.4 (93.3–99.5) | 0.025 |

DDFS, distant disease-free survival; EFS, event-free survival; HR, hazard ratio; IBCFS, invasive breast cancer-free survival; iDFS, invasive disease-free survival; OS, overall survival; SD, standard deviation. EFS and OS were measured from randomization among the 309 patients with a valid baseline RISK score; iDFS, IBCFS and DDFS were measured from the date of surgery among the 290 patients who underwent protocol-defined surgery. Patients analyzed excludes those with no recorded follow-up time (18 for EFS, 19 for OS, and 7 of the 290 for the surgery-anchored endpoints). The two blocks are different analyses of the same endpoint: the left-hand block uses the continuous RISK score, the right-hand block the prespecified low- and high-risk cutoff. The P values are therefore not interchangeable. DDFS events comprise distant recurrence or death from any cause; the 17 DDFS events consist of 15 distant recurrences and 2 deaths without prior distant recurrence. Events by risk group (low vs high): EFS 13/24; iDFS 10/16; IBCFS 9/13; DDFS 4/13; OS 1/7. Kaplan-Meier estimates with log-transformed 95% confidence intervals.

Additional analyses for iDFS, IBCFS, DDFS, and overall survival (OS) are shown in **Table 3** and **Figure 3**. Of the 15 distant recurrences observed, 12 (80.0%) occurred in the high-risk group. For OS, the HR per 1-SD increase in the RISK score was 1.72 (95% CI, 0.96 to 3.11; P=0.070; Harrell’s C-index, 0.73). Eight deaths occurred, one in the low-risk group and seven in the high-risk group.

The prognostic value of the RISK score was further evaluated according to pathologic response to neoadjuvant therapy. Among the 186 patients who did not achieve pCR (167 with pathologically documented residual disease at protocol-defined surgery and 19 who did not undergo protocol-defined surgery and were therefore classified as non-pCR), the RISK score was associated with EFS (HR per 1-SD increase, 1.65; 95% CI, 1.22 to 2.23; P=0.001; 31 events). Analyses of iDFS, IBCFS, and DDFS were restricted to the 167 patients in this subgroup who underwent definitive surgery and showed HRs per 1-SD increase in the RISK score of 1.21 (95% CI, 0.80 to 1.84; P=0.370; 20 events), 1.25 (95% CI, 0.81 to 1.93; P=0.307; 18 events), and 1.37 (95% CI, 0.88 to 2.12; P=0.158; 16 events), respectively. Among the 123 patients who achieved pCR, the number of events was low across endpoints (EFS, 6 events; iDFS, 6 events; IBCFS, 4 events; DDFS, 1 event), with no statistically significant associations observed between the RISK score and clinical outcomes. RISK-by-pCR interaction tests were not statistically significant for EFS (P=0.48), iDFS (P=0.94), or IBCFS (P=0.19). Interactions for DDFS and OS could not be meaningfully assessed, because only one distant event and no deaths occurred among patients who achieved pCR. RISK estimates from baseline biopsies and surgical specimens were concordant and showed generally consistent associations with outcomes (**Table S2**).

## DISCUSSION

In this retrospective analysis of two-phase II trials, we found that the Ataraxis Breast multimodal AI platform provided both predictive and prognostic information in HER2+ EBC evaluated in the context of CT de-escalation strategies. The NEO model was consistently associated with pCR in both PHERGain and PHERGain-2, while the RISK model identified patients with significantly different EFS outcomes in PHERGain. These findings support further prospective evaluation of multimodal AI applied to routine histopathology in HER2+ EBC.

The ability to identify patients most likely to achieve pCR with HER2-targeted therapy remains a major unmet need in treatment de-escalation. Higher NEO scores were associated with pCR in both trials, and NEO significantly improved models based on clinicopathological variables, whereas adding these variables to NEO provided no significant improvement. Although the AUCs indicated modest overall discrimination, the prespecified NEO cutoffs identified substantial differences in pCR rates between the low- and high-probability groups in both trials. However, separation between the intermediate- and high-probability groups was more limited in PHERGain (41.3% vs 48.3%) and PHERGain-2 (61.6% vs 70.1%), suggesting that the prespecified NEO cutoffs may provide greater discrimination in identifying patients with a low probability of pCR than in distinguishing between intermediate and high probabilities of response. This pattern may be clinically relevant for treatment de-escalation, as NEO could be particularly useful for identifying patients unlikely to achieve pCR and who may therefore be less suitable for chemotherapy-free strategies, rather than for distinguishing between patients with intermediate and high probabilities of response.

In exploratory PHERGain subgroup analyses, the NEO score demonstrated greater discrimination in Group A (upfront TCHP) than in Group B (18F-FDG-PET-adapted treatment), with pCR rates ranging from 25.0% to 73.9% versus 18.8% to 41.9%, respectively. Analyses by PET response were limited by small sample sizes, and no significant NEO-by-PET interaction was detected. The attenuated association in Group B may reflect treatment escalation among PET non-responders.

The RISK score was prognostic for EFS in PHERGain, with most distant recurrences and deaths occurring in the high-risk group. However, associations between the continuous score and secondary outcomes were not statistically significant and should be interpreted cautiously given the limited number of events.

Our findings complement prior efforts to develop biomarkers for treatment personalization in HER2+ EBC. Notably, HER2DX has also been evaluated in PHERGain, demonstrating a significant association with pCR and providing prognostic information on recurrence risk ^7^. While HER2DX integrates gene-expression profiles reflecting immune response, luminal differentiation, proliferation, and HER2 amplicon biology with clinical features, ATX derives information from AI-based analysis of routine HCE whole-slide images combined with clinical variables. Thus, these approaches interrogate tumor biology through distinct modalities and may provide complementary perspectives on treatment sensitivity and prognosis. Similarly, circulating tumor DNA (ctDNA) analyses from PHERGain provide a dynamic, treatment-dependent assessment of molecular response, with persistent ctDNA detection and failure to achieve early ctDNA clearance associated with lower pCR rates and worse long-term outcomes. Collectively, these findings highlight the potential of genomic, digital pathology, and circulating biomarkers to capture different dimensions of tumor biology and treatment response and provide a rationale for future studies evaluating their potential integration in treatment de-escalation strategies.

Importantly, although retrospective biomarker analyses embedded within prospective clinical trials provide a robust framework for assessing biomarker performance, further evidence of clinical utility will be needed before these tools can be used to guide treatment de-escalation in routine practice. Future studies should determine whether NEO- and RISK-informed treatment decisions can safely reduce treatment burden and toxicity while preserving oncologic outcomes.

This study has several limitations. The biomarker analyses were retrospective, although specimens and data were prospectively collected within two phase II trials. NEO discrimination was modest, and prognostic analyses were restricted to PHERGain because time-to-event data were not mature for PHERGain-2. Although the associations observed were consistent across cohorts, the discriminative performance was moderate, suggesting that the model should be viewed as a complementary tool rather than a standalone predictor and warranting further validation in additional datasets. The small number of recurrence and survival events also limited the precision of secondary prognostic analyses.

Despite these limitations, AI-based analysis of routine HCE slides could offer a potentially simple, rapid, scalable, and resource-efficient approach to treatment personalization. The predictive and prognostic information provided by NEO and RISK scores supports further investigation of multimodal AI as a tool to refine patient selection for chemotherapy de-escalation in HER2+ EBC. Future studies should determine the clinical utility of integrating these models with clinicopathologic factors and other molecular and dynamic response biomarkers, with the ultimate goal of enabling more precise and individualized treatment strategies while safely reducing treatment burden.

## DATA AVAILABILITY STATEMENT

Raw sequencing data generated and analyzed in this study are not publicly available owing to proprietary and commercial confidentiality constraints. De-identified individual participant data are available under controlled access. Access is restricted because these are clinical trial data subject to the informed consent obtained from participants and to the sponsor’s legal and contractual obligations. Requests should be directed to the corresponding author (Dr Antonio Llombart) and are subject to review and approval by the PHERGain and PHERGain-2 trial management group, including qualified statisticians. Access requires a detailed research proposal and a signed data-sharing agreement with the study sponsor. Requests will be answered within 30 days. Data will be available from 1 month to 5 years after publication of this article. All data will be anonymized in accordance with applicable laws and regulations.

## ETHICS APPROVAL

The PHERGain study (MedOPP096-MO39229; EudraCT 2016-002676-27) received favorable approval from the Research Ethics Committee with Medicines (CEIm) of Hospital Arnau de Vilanova-Llíria, Valencia, Spain. The PHERGain-2 study protocol (MedOPP293; EudraCT 2020-003205-66; CEIC-2436) received favorable approval from the Research Ethics Committee with Medicines (CEIm) of Hospital Universitari Arnau de Vilanova (Lleida, Spain).

## ACKNOWLEDGMENTS

Hoffmann-La Roche funded PHERGain and PHERGain-2 trials and ATARAXIS AI funded the multimodal AI analysis. We wish to thank the patients who kindly participated in our study and their families. We also acknowledge the Investigators and their teams.

