## Appendix for "Multimodal Artificial Intelligence Predicts Pathological Response and Prognosis in HER2-Positive Early Breast Cancer Treated with Chemotherapy De-escalation Strategies: Analyses from PHERGain and PHERGain-2 Trials"

### Table S1. Diagnostic performance NEO score association for pCR

| Term in multivariable model | Adjusted OR (95% CI) | P |
|---|---|---|
| **PHERGain (n = 302; adjusted for treatment group + hormone receptor status)** | | |
| *Continuous NEO score* | | |
| **NEO score, per 0.1-unit increase** | **1.24 (1.02–1.52)** | **0.033** |
| Treatment arm: adaptive vs control | 0.40 (0.21–0.74) | 0.004 |
| HR status: positive vs negative | 0.92 (0.46–1.82) | 0.801 |
| *Prespecified NEO probability groups* | | |
| **NEO medium *vs* low** | **2.64 (1.25–6.04)** | **0.015** |
| **NEO high *vs* low** | **2.77 (1.13–7.17)** | **0.030** |
| Treatment arm: adaptive vs control | 0.42 (0.22–0.77) | 0.006 |
| HR status: positive vs negative | 0.69 (0.36–1.32) | 0.269 |
| **PHERGain-2 (n = 383; adjusted for hormone receptor status)** | | |
| *Continuous NEO score* | | |
| **NEO score, per 0.1-unit increase** | **1.43 (1.18–1.74)** | **<0.001** |
| HR status: positive vs negative | 1.61 (0.88–3.00) | 0.13 |
| *Prespecified NEO probability groups* | | |
| **NEO medium *vs* low** | **3.99 (2.01–8.31)** | **<0.001** |
| **NEO high *vs* low** | **6.85 (2.91–16.84)** | **<0.001** |
| HR status: positive vs negative | 1.34 (0.75–2.42) | 0.329 |

CI, confidence interval; NEO, Ataraxis Breast NEO score; OR, odds ratio; pCR, pathological complete response. Logistic regression with pCR (ypT0/isN0) as the outcome. Odds ratios are per 0.1-unit increase in the continuous NEO score, or versus the low-probability group. Confidence intervals are profile likelihood. Models adjust for treatment group and hormone receptor status in PHERGain, and for hormone receptor status in PHERGain-2. The PHERGain multivariable population is the 302 patients of the 306 NEO-analyzable set with treatment group and hormone receptor status both recorded.

### Table S2. Association between Ataraxis Breast NEO risk score and long-term clinical outcomes at baseline and surgery

| Endpoint | n | Events | HR baseline (95% CI), p | HR surgery (95% CI), P |
|---|---|---|---|---|
| iDFS | 225 | 23 | 1.18 (0.80–1.74), 0.402 | 1.33 (0.83–2.12), 0.231 |
| IBCFS | 225 | 19 | 1.04 (0.67–1.63), 0.847 | 1.19 (0.69–2.05), 0.522 |
| DDFS | 225 | 14 | 1.33 (0.83–2.14), 0.240 | 1.62 (0.95–2.77), 0.076 |
| EFS | 227 | 29 | 1.53 (1.11–2.11), 0.010 | 1.74 (1.21–2.50), 0.003 |
| OS | 227 | 5 | 2.01 (0.99–4.11), 0.054 | 2.15 (1.00–4.62), 0.049 |

### Table S3. Incremental value of Ataraxis NEO over clinicopathological variables for the prediction of pathological complete response.

| Specification | AUC (corrected) | | | ΔAUC | Likelihood ratio test | |
|---|---|---|---|---|---|---|
| | NEO alone | Clinical | Clinical + NEO | NEO added | NEO added to clinical model | Clinical added to NEO |
| **PHERGain (n = 306)** | | | | | | |
| Cohort-specific<br>*age, cT, cN, grade, HR* | 0.613 | 0.567 | 0.611 | +0.043 | $\chi^2$ = 11.7 (1 df)<br>P < 0.001 | $\chi^2$ = 12.8 (8 df)<br>P = 0.12 |
| Common model<br>*age, cT, grade, HR* | 0.613 | 0.565 | 0.614 | +0.049 | $\chi^2$ = 12.1 (1 df)<br>P < 0.001 | $\chi^2$ = 11.8 (6 df)<br>P = 0.066 |
| **PHERGain-2 (n = 378)** | | | | | | |
| Common model<br>*age, cT, grade, HR* | 0.617 | 0.518 | 0.592 | +0.073 | $\chi^2$ = 12.4 (1 df)<br>P < 0.001 | $\chi^2$ = 4.7 (5 df)<br>P = 0.45 |

Clinical models included age, clinical tumor stage, histological grade, and hormone receptor status; clinical nodal stage was additionally included for PHERGain. Analyses used patients with complete covariate data. AUC, area under the receiver operating characteristic curve; cT, clinical tumour stage; cN, clinical nodal stage; HR, hormone receptor. All AUCs are optimism-corrected by Harrell's bootstrap. ΔAUC is the increment from adding NEO to the clinical model and equals the difference between the corresponding corrected columns. Added value was assessed by likelihood ratio. The two tests address different hypotheses: that NEO adds nothing to the clinical model, and that the clinical covariates add nothing to NEO. Analyses are restricted to patients with complete covariate data. In PHERGain-2 this excludes five patients with missing histological grade.

### Figure S1. ROC curves for the continuous NEO model predicting pCR.

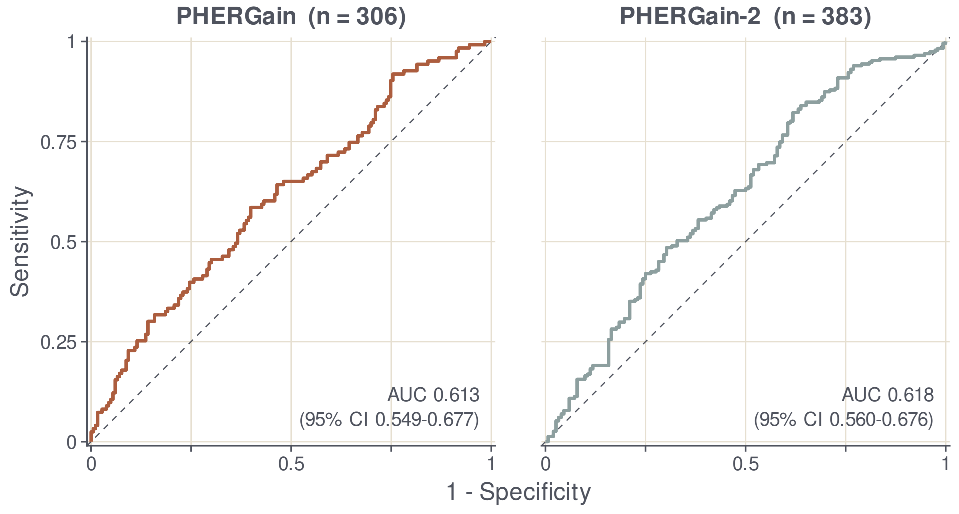


### Supplementary Methods

*Ataraxis Breast NEO model*

The Ataraxis Breast NEO model is a two-stage multimodal artificial intelligence framework developed to predict pathological complete response (pCR) using digitized hematoxylin and eosin (H&E) whole-slide images and clinical variables. Pathology foundation models (ViT-g, 1.1B parameters; ViT-L, 300M parameters) generated embeddings from non-overlapping 224×224-pixel patches extracted at 0.5 μm/pixel from pretreatment core-biopsy slides.

A gated-attention multiple-instance learning (MIL) framework aggregated patch-level representations and inferred the expression of 14,773 genes. The gene-expression inference module was trained using 7,054 The Cancer Genome Atlas (TCGA) cases and externally evaluated using Clinical Proteomic Tumor Analysis Consortium (CPTAC) datasets. Inferred gene-expression profiles were averaged at the patient level, restricted to a prespecified panel of 2,427 genes, and reduced to five independent component analysis (ICA) features.

The resulting molecular representations were integrated with eight baseline clinicopathologic variables (ER, PR, HER2, clinical T stage, clinical N stage, age, ductal indicator, and lobular indicator) using an ensemble consisting of an explainable boosting machine and a tabular residual network (ResNet). Model development included 1,080 patients across five cohorts using multiple-source cross-validation and evaluation in 1,412 patients across nine independent cohorts.

Prespecified thresholds categorized NEO scores into low (<0.30), medium (0.30–0.55), and high (>0.55) probability of pCR.

*Ataraxis Breast RISK model*

The Ataraxis Breast RISK model is a multimodal artificial intelligence framework developed to estimate 5-year recurrence risk using H&E whole-slide images and clinical variables. A pathology foundation model (DINOv2 ViT-g, 1.1B parameters) generated embeddings from non-overlapping image patches extracted at 20× magnification (0.5 μm/pixel).

Patch-level representations were aggregated using gated-attention MIL to infer expression levels for 14,773 genes. The gene-expression inference module was trained using TCGA data and externally evaluated using CPTAC datasets. Patient-level inferred expression profiles were restricted to the 1,000 genes with the highest Kendall's τ in the TCGA validation set and reduced to 200 principal component analysis (PCA) features.

The survival prediction framework combined inferred gene-expression features, image-derived embeddings, and clinicopathologic variables through an ensemble of Cox proportional hazards models, discrete-time survival models, and tabular residual networks. Clinical variables included age, ER, PR, HER2, primary and secondary T stage, primary and secondary N stage, and indicators of invasive ductal and invasive lobular histology.

Model development included 9,947 patients from 16 institutions, internal validation in 3,693 patients, and independent evaluation in 5,554 patients from nine institutions. Models were trained to predict recurrence at any site and calibrated for 5-year recurrence risk using beta calibration.

A prespecified threshold classified patients as low risk ($<0.20$) or high risk ($\geq 0.20$) for 5-year recurrence.
